# Impact of the COVID-19 pandemic and response on the utilisation of health services during the first wave in Kinshasa, the Democratic Republic of the Congo

**DOI:** 10.1101/2021.04.08.21255096

**Authors:** Celestin Hategeka, Simone Elyse Carter, Faustin Mukalenge Chenge, Eric Nyambu Katanga, Grégoire Lurton, Serge Ma-Nitu Mayaka, Dieudonné Kazadi Mwamba, Esther van Kleef, Veerle Vanlerberghe, Karen Ann Grépin

## Abstract

**Introduction:** Health service use among the general public can decline during infectious disease outbreaks and has been predicted among low and middle-income countries during the COVID-19 pandemic. In March 2020, the government of the Democratic Republic of the Congo (DRC) implemented public health measures across Kinshasa, including strict lockdown measures in the Gombe health zone, to mitigate impact of the pandemic.

**Methods:** Using data from the Health Management Information System (January 2018 - December 2020), we evaluated the impact of the pandemic on the use of essential health services (total visits, maternal health, vaccinations, visits for common infectious diseases, and diagnosis of non-communicable diseases) using interrupted time series with mixed effects segmented Poisson regression models during the first wave of the pandemic. Analyses were stratified by age, sex, health facility, and neighbourhood.

**Results:** Health service use dropped rapidly following the start of the pandemic and ranged from 16% for hypertension diagnoses to 39% for diabetes diagnoses. However, reductions were highly concentrated in Gombe (81% decline in total visits) relative to health zones without lockdown. When the lockdown was lifted, total visits, visits for infectious diseases, and diagnoses for non-communicable diseases increased approximately two-fold. Hospitals were more affected than health centres. Overall, the use of maternal health services and vaccinations was not significantly affected.

**Conclusion:** The COVID-19 pandemic resulted in important reductions in health service utilisation in Kinshasa, particularly Gombe. Lifting of lockdown led to a rebound in the level of health service use but it remained lower than pre-pandemic levels.

**Summary Box:** *What is already known about this subject:*

- Substantial declines in the use of health services among the general public have been well-documented during previous outbreaks of infectious diseases.
- Modelled studies predicted substantial increases in morbidity and mortality in many low- and middle-income countries (LMICs) mainly due to expected declines in the use of health services among the general public.
- Only a small number of studies have so far evaluated the impact of the COVID-19 pandemic on the use of health services in LMICs and none have also evaluated both the implementation and lifting of lockdown measures.

*What are the new findings:*

- This study found that overall use of health services declined in Kinshasa but was most pronounced in the Gombe health zone which was subject to strict lockdown measures.
- Some health services were more affected than others, most notably visits and tests for malaria and visits for new diagnoses of non-communicable diseases. Maternal and child health services were relatively unaffected.
- When the lockdown measures were lifted, health service utilization rebounded but remained at levels lower than those observed pre-pandemic.

*What do the new findings imply:*

- The COVID-19 pandemic has likely had important effects on the use of health services among the general public throughout LMICs. However, evidence from Kinshasa suggests the effects may not be as widespread as previously assumed.
- The impact of strict social distancing measures needs on COVID-19 outcomes needs to be weighed off against the potential population-level health effects of these policies in various international contexts.

## INTRODUCTION

At the outset of the COVID-19 pandemic, it was predicted that Sub-Saharan Africa (SSA) was vulnerable to record large numbers of cases and deaths.^1^ To date, however, the region has been relatively less affected – through March 22, 2021, it accounted for only approximately 2.5% of globally confirmed cases and a lower proportion of deaths.^2^ It is unclear, however, to what extent these levels may be explained by lower testing rates, less severe clinical presentation, or other factors.

Studies have shown that outbreaks, as well as governmental response to outbreaks, can lead to important unintentional secondary health effects - or “collateral damage” - mainly the result of reduced utilization of health services for other conditions. For example, a systematic review of the West African Ebola Virus Disease (EVD) outbreak found an 18% decline in the overall use of health services.^3^ Some health services were more severely affected: reductions of 80% in facility deliveries, 40% reductions in malaria admissions, and important reductions in immunizations were all documented.^4^ Modelling studies have suggested that mortality from non-Ebola conditions was potentially as large as the direct effects of EVD.^5–8^ Many factors likely led to decreased use of health services, including interruptions in treatment protocols,^9–11^ health worker mortality,^12^ as well as fear and a lack of trust in the health system.^13,14^ Persistent reductions in health service utilization were also observed in the aftermath of the West African EVD outbreak.^15–17^

There are concerns that such patterns will be repeated during COVID-19, especially in SSA where sizeable increases in health service utilization over the past few decades could be erased. Early modelled studies predicted that the pandemic could exert a devastating toll on health service utilization and mortality.^18,19^ Early predictions of the potential impact of health service disruptions on the burden of malaria in SSA suggested that under certain scenarios, malaria deaths could double in 2020 relative to 2018.^20,21^ Another study estimated that drops in the coverage of maternal and child health interventions could lead to substantial additional deaths in low and middle-income countries (LMICs).^18^

While modelled studies provide useful insights, their ability to accurately predict outcomes depends on the data available, the models used, and the parameter assumptions made, all of which can be highly uncertain or incomplete, especially at the start of an outbreak.^22^ The situation may be even more challenging in LMIC settings, where data is more limited.

To date, only a few observational studies have investigated the impact of the pandemic on the use of health services in LMICs. In South Africa, one study found that lockdowns were associated with a large and significant decline in the use of health services among children but not adults.^23^ In Karachi, Pakistan, a lockdown introduced in late March was associated with a more than 50% decline in routine immunizations.^24^ In Nepal, institutional delivery rates decreased by more than half during a lockdown period.^25^ No studies, to our knowledge, have yet investigated the impact of the lifting of such lockdown policies.

In early 2020, the Democratic Republic of the Congo (DRC) was already dealing with many competing outbreaks, including large-scale outbreaks of EVD and measles.^26^ Given its large population, densely populated cities, and weak health system, it was considered to be highly vulnerable to COVID-19.^1^ As of March 22, 2021, however, the country had confirmed only 27,552 cases and 726 deaths.^2^

Policymakers need real-time data and evidence to inform decisions during outbreaks.^27^ Data from routine health information systems (RHIS) are now widely available in LMICs and have been used conduct similar analyses during other infectious disease outbreaks.^17,28,29^ In this study, we use RHIS data to evaluate the impact of COVID-19 and its related response measures on the use of health services in Kinshasa during the first wave of the pandemic (March-September) to provide insights to inform the ongoing response and future infectious disease outbreaks.

## METHODS

### Context

With a population of over 14 million, Kinshasa is one of the largest and most densely populated cities in Africa. The DRC health system is organized into health zones, which are further disaggregated into health areas. Each health zone should have at least one hospital, while each health area should have at least one health centre. Currently, Kinshasa has 851 health centres and 121 hospitals—some of which were designated COVID-19 treatment centres (Figure 1). There is also an active private sector, which plays an important complementary role in delivering health services.^30^

**Figure 1:**
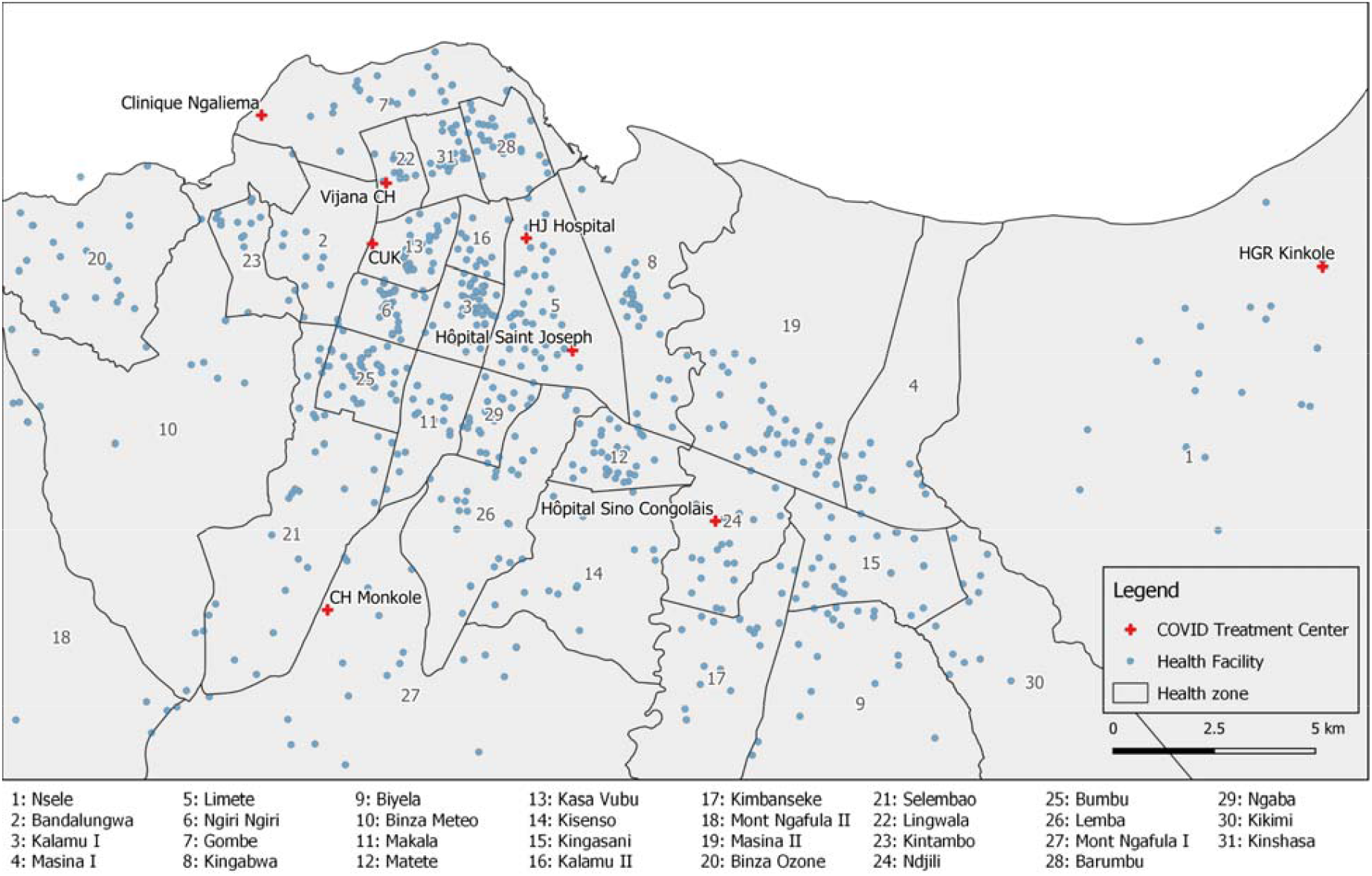
A map of Kinshasa, with health zones outlined and showing 8 health facilities, initially identified as centres for COVID- 19 cases treatment and hospitalization (March-April 2020).

The first case of COVID-19 in the DRC was identified on March 10, 2020.^31^ Over the next few weeks, the government introduced an outbreak management and control plan including a series of public health measures aimed at reducing transmission of the virus. On April 6, 2020, the neighbourhood of Gombe, at the time known as the epicentre of the epidemic, was locked down, which closed stores and restricted all non-essential travel in and out of the neighbourhood and limited all movement within the area to only essential travel. The lockdown was partially lifted on April 21, allowing residents to purchase food and other essentials, but remained in place until June 29. There was no lockdown outside of the Gombe health zone. Figure 2 provides an overview of the confirmed cases of COVID-19 in Kinshasa and other DRC provinces.

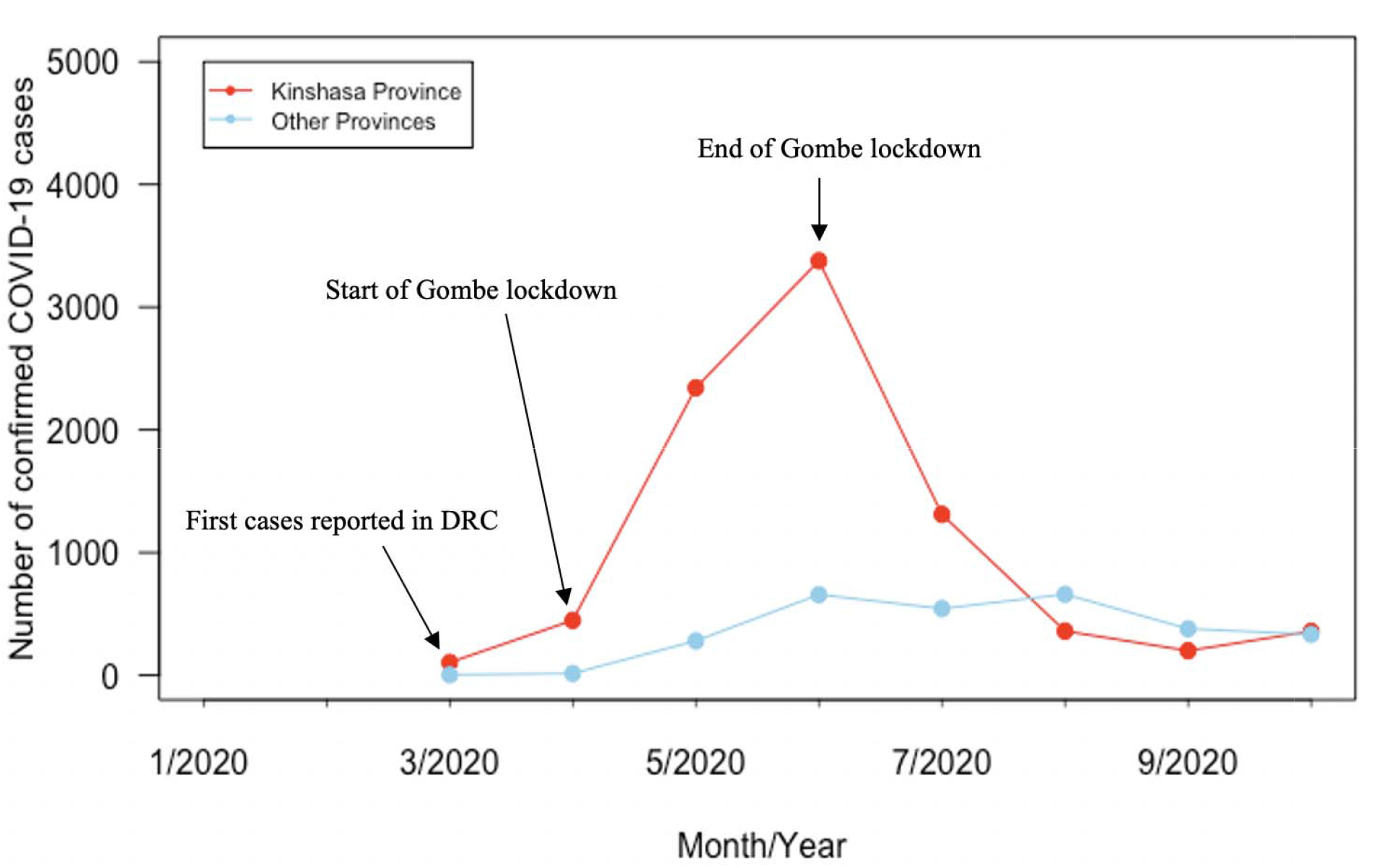

### Outcomes

We evaluated the impact of COVID-19 on 14 indicators of health service utilization:

- *overall*: total clinic visits;
- *common infectious diseases*: uncomplicated pneumonia cases diagnosed; uncomplicated diarrhea cases diagnosed; uncomplicated malaria cases diagnosed, and rapid diagnostic tests (RDT) for malaria conducted;
- *maternal health*: first antenatal care visit (ANC 1), institutional deliveries, postnatal care within 6 days of birth (PNC 2);
- *vaccinations*: first doses of Diphtheria-Tetanus-Pertussis vaccine (DTP) administered, BCG, Oral Polio Vaccine (OPV) 1, and the first dose of Pneumococcal Conjugate Vaccine (PCV-13);
- *Non-communicable diseases (NCDs):* Hypertension and diabetes cases newly diagnosed.

These indicators were selected because they accounted for the majority of primary care services provided by health facilities as well as those we believed could be influenced by the pandemic (see Appendix Table 1) as well as indicators with high completeness reporting rates, except for the pneumonia and the NCD indicators, which had reporting rates less than 60% but were still included to provide a more comprehensive picture of health service utilization.

### Analytical strategy

We used ITS analyses to assess the impact of the onset of the pandemic and the government response measures, using monthly time series data, while controlling for secular trends in the outcomes.^34,35^ As March 2020 was partially exposed to both the pandemic and the Gombe lockdown, we defined the start of both events as of April 2020 and the Gombe lockdown period as April-June 2020. As baseline rates in health service volume vary across health facilities, we employed segmented quasi-Poisson mixed-effects models, with health facility catchment population as an offset to estimate the impact on each indicator immediately following the start of the pandemic or the Gombe lockdown (level change) and over time (trend change) (see Statistical Appendix). All our models were also adjusted for seasonality. Additionally, models for total clinic visits and visits for common infectious diseases included a dummy variable to adjust for an unrelated pneumonia outbreak that took place in Kinshasa from December 2019 to February 2020. We also provide results from analyses that were not adjusted for the pneumonia outbreak in the Appendix as a sensitivity analysis (see Appendix Table 2).

We only included health centres and hospitals in our analysis. We excluded health posts, which provide largely health promotion and community health services, and private health facilities because their reporting rates are limited. For each indicator, we included health facilities that had a reporting rate of at least 25% before COVID-19 (January 2018-February 2020) and after the onset of the pandemic (March-December 2020). We defined outliers for each indicator as any observation exceeding seven standard deviations from the meantime trend estimated using facility-level local regression, which were subsequently treated as missing observations. Facilities with consecutive missing observations for a specific indicator were excluded from our analyses. Because of these inclusion criteria, the number of facilities included in our analyses varied by indicator (see Appendix Table 3). Missing data were imputed using seasonally decomposed missing value imputation, accounting for seasonal patterns in the service utilization time series data. We also performed sensitivity analyses using complete case analyses—i.e., analyses that include facilities that had complete reporting or no missing values during the study period. Findings from the complete case analyses were largely comparable to the reported results (see Appendix Table 5).

We also conducted subgroup analyses. First, wherever possible we stratified our analyses by the Gombe versus the remaining 34 health zones to estimate the additional impact of the lockdown versus COVID-19 alone. For the Gombe health zone, we also ran models that included segments (level and trend change) for the lockdown (April-June 2020) and post lockdown (July-December 2020) periods, allowing us to also estimate the impact of stopping the policy (see Statistical Appendix). Second, we conducted additional analyses to investigate whether the pandemic had a differential impact on different groups, specifically we stratified our sample by gender, age, and health facility type wherever feasible. All analyses were conducted using R version 4.0.2. We report parameter estimates using the incidence rate ratio (IRR) and related 95% confidence interval (CI).

### Ethical statement

We used a research protocol that had been approved by the Ethics Committees at Wilfrid Laurier University (Canada) and Kinshasa School of Public Health (DRC). We also obtained authorization from the Ministry of Public Health to utilise these data to evaluate the impact of the pandemic on health service utilization.

## RESULTS

Our analyses included 9,158,657 total clinic visits, 3,282,890 visits for infectious diseases (malaria, diarrhea, and pneumonia), 256,405 new diagnoses of noncommunicable diseases (diabetes and hypertension), and 3,467,713 maternal and child health services (first antenatal care visits, institutional deliveries, second postnatal care visits, and vaccinations) between January 2018 and December 2020 (see Appendix Table 4).

### Overall

We found that the overall use of health service decreased following the start of the pandemic (Table 1). Rates of total clinic visits immediately decreased by 25% (IRR: 0.75, 95% CI: 0.73—0.75) relative to the pre-pandemic period (Table 1 and Figure 3A). However, rates of total clinic visits increased marginally after the initial decline, but significantly, over time relative to the trend that would have been expected without COVID-19 (IRR: 1.01, 95% CI: 1.00—1.02). Results stratified by areas with and without lockdown showed that the policy resulted in an immediate decrease of over 80% in rates of all clinic visits in Gombe (IRR: 0.19, 95% CI: 0.12—0.32); however, the rates of clinic visits immediately increased by more than two-fold when the lockdown was lifted (IRR: 2.50, 95% CI: 1.81—3.45), but remained lower than pre-pandemic rates by approximately 16% (Table 2. Figure 3B and Appendix Table 6). In contrast, health zones without lockdown had relatively smaller declines of about only 15% (IRR: 0.84, 95% CI: 0.81—0.86) (Table 2 and Figure 3B). Analyses stratified by gender showed that there was a similar immediate reduction in the clinic visits in both female and male patients (Table 1 and Appendix Figure 1). Similarly, decreases in service use were very similar among under five-year patients compared with those over five years of age (Table 1). However, immediate reductions in the total clinic visits were substantially higher in hospitals (IRR: 0.57, 95% CI: 0.53—0.62) than in health centres (IRR: 0.86, 95% CI: 0.84—0.89) (Table 1 and Appendix Figure 2). Additional analysis stratified by COVID-19 and non-COVID-19 hospitals showed that reductions in service use were even higher in COVID-19 hospitals (Table 1).

**Table 1.** Parameter estimates of the overall effect of COVID-19 on rates of total clinic visits, visits for common infectious diseases and non-communicable diseases, and maternal and child health services in Kinshasa, DRC.

| Indicators | Level change |  | Trend change |  |
| --- | --- | --- | --- | --- |
|  | IRR (95% CI) | p-value | IRR (95% CI) | p-value |
| **Overall visits and by sex, age and facility tier** |  |  |  |  |
| Total clinic visits | 0.75 (0.73, 0.75) | <0.001 | 1.01 (1.00, 1.02) | <0.001 |
| • Female | 0.76 (0.73, 0.79) | <0.001 | 1.01 (1.00, 1.02) | <0.001 |
| • Male | 0.74 (0.71, 0.77) | <0.001 | 1.01 (1.00, 1.01) | <0.001 |
| • Under five years | 0.75 (0.73, 0.77) | <0.001 | 1.00 (1.00, 1.01) | 0.01 |
| • Five years and over | 0.75 (0.73, 0.77) | <0.001 | 1.01 (1.00, 1.02) | <0.001 |
| • Health centers | 0.86 (0.84, 0.89) | <0.001 | 1.01 (1.00, 1.01) | <0.001 |
| • Hospitals | 0.57 (0.53, 0.62) | <0.001 | 1.02 (1.00, 1.03) | 0.009 |
| • COVID-19 Hospitals | 0.48 (0.31, 0.74) | <0.001 | 1.03 (0.97, 1.11) | 0.27 |
| • Non-COVID-19 hospitals | 0.58 (0.53, 0.63) | <0.001 | 1.01 (1.00, 1.02) | 0.02 |
| **Common IDs** |  |  |  |  |
| Malaria | 0.76 (0.73, 0.79) | <0.001 | 1.01 (1.00, 1.02) | 0.001 |
| RDT | 0.70 (0.67, 0.73) | <0.001 | 1.01 (1.01, 1.02) | <0.001 |
| Diarrhea | 0.74 (0.70, 0.78) | <0.001 | 1.01 (1.00, 1.02) | <0.001 |
| Pneumonia | 0.70 (0.65, 0.74) | <0.001 | 1.03 (1.02, 1.04) | <0.001 |
| **NCDs** |  |  |  |  |
| Diabetes | 0.61 (0.55, 0.66) | <0.001 | 1.03 (1.01, 1.04) | <0.001 |
| Hypertension | 0.84 (0.79, 0.89) | <0.001 | 1.00 (0.99, 1.02) | 0.15 |
| **Maternal health** |  |  |  |  |
| ANC 1 | 1.04 (1.01, 1.07) | 0.007 | 0.99 (0.99, 1.00) | 0.71 |
| Institutional deliveries | 1.08 (1.05, 1.11) | <0.001 | 1.00 (0.99, 1.01) | 0.32 |
| PNC 2 | 1.07 (1.03, 1.11) | <0.001 | 1.00 (0.99, 1.01) | 0.16 |
| **Vaccinations** |  |  |  |  |
| DTP1 | 1.01 (0.98, 1.04) | 0.40 | 1.00 (0.99, 1.01) | 0.26 |
| BCG | 0.95 (0.90, 1.01) | 0.08 | 1.01 (1.00, 1.02) | 0.001 |
| OPV | 1.04 (1.01, 1.08) | 0.004 | 1.00 (1.00, 1.01) | 0.005 |
| PCV-13 | 1.00 (0.97, 1.04) | 0.60 | 1.00 (0.99, 1.01) | 0.47 |

IRR: incidence rate ratio; CI: confidence interval; IDs, infectious diseases; RDT, rapid diagnostic test for malaria; NCDs, non-communicable diseases; ANC, antenatal care; PNC, postnatal care; DTP: diphtheria, pertussis, and tetanus vaccine; BCG, Bacillus Calmette–Guérin vaccine; OPV, Oral poliovirus vaccine; PCV-13, Pneumococcal Conjugate Vaccine. Parameter estimates in Table 1 are from mixed effects segmented regression models comparing the period before COVID-19 (January 2018–February 2020) and after the onset of COVID-19 (April–December 2020) in DRC.

**Table 2.**
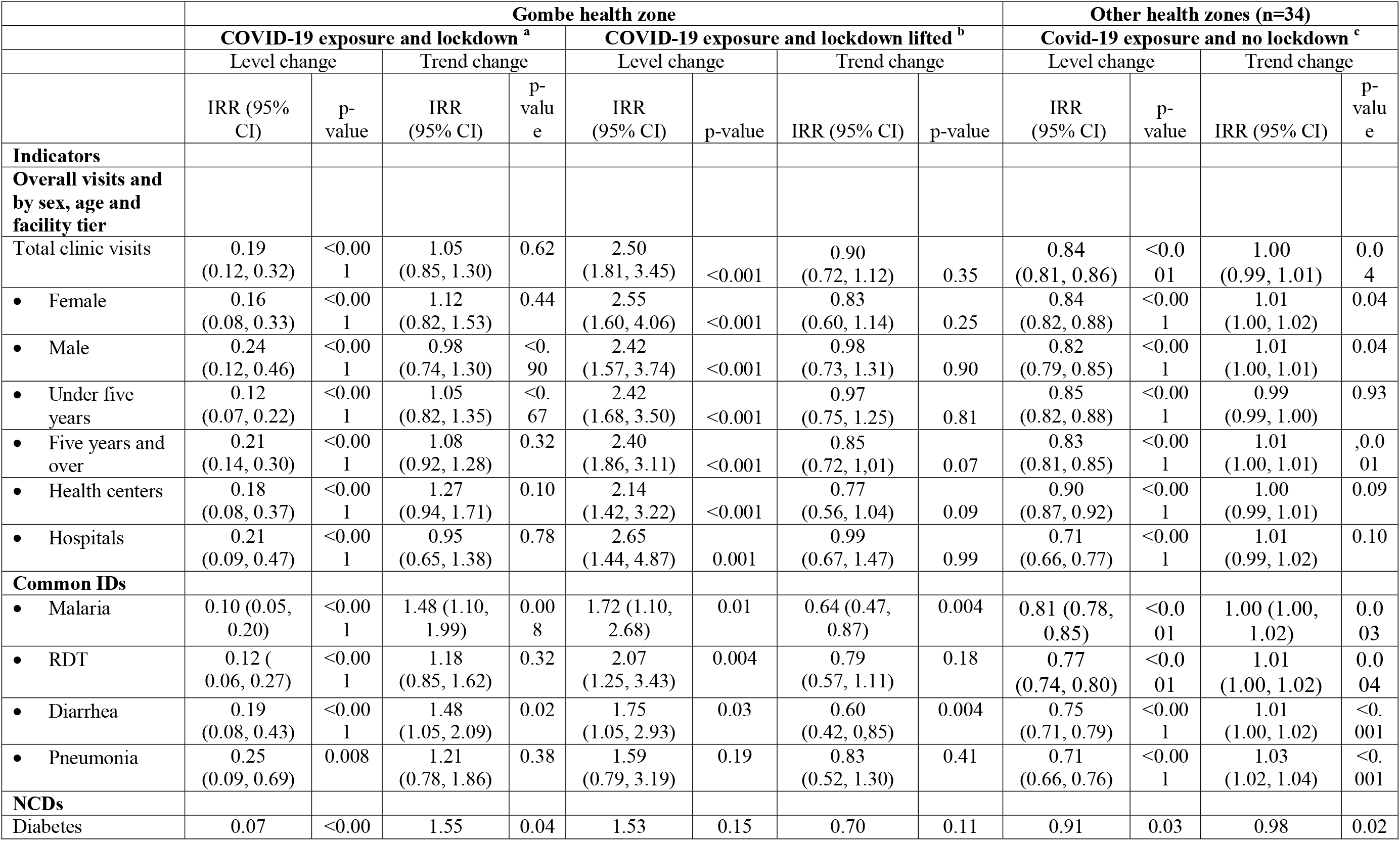
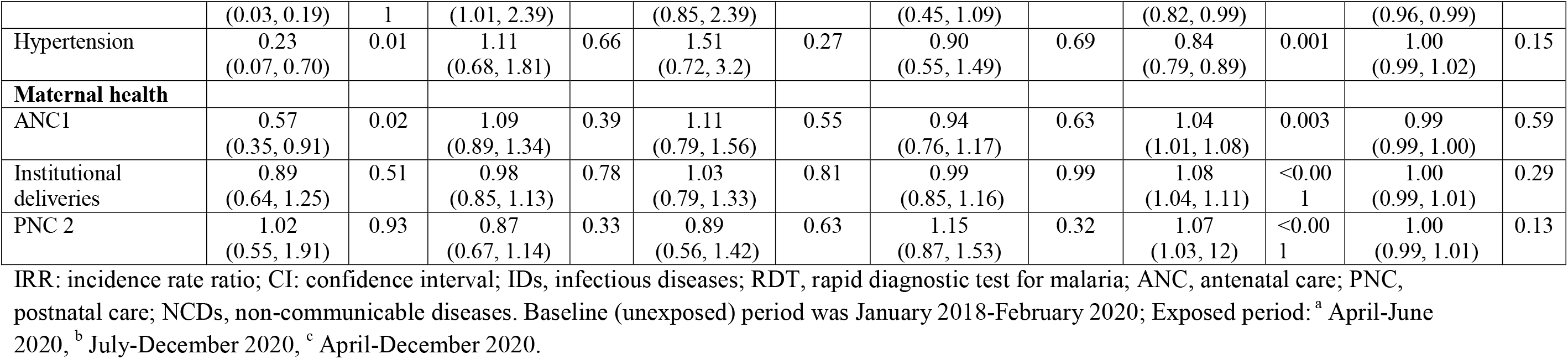
Parameter estimates of the effect COVID-19 and the lockdown policy on rates of total clinic visits, visits for common infectious diseases and non-communicable diseases, and maternal and child health services in Kinshasa, DRC.

**Figure 3.**
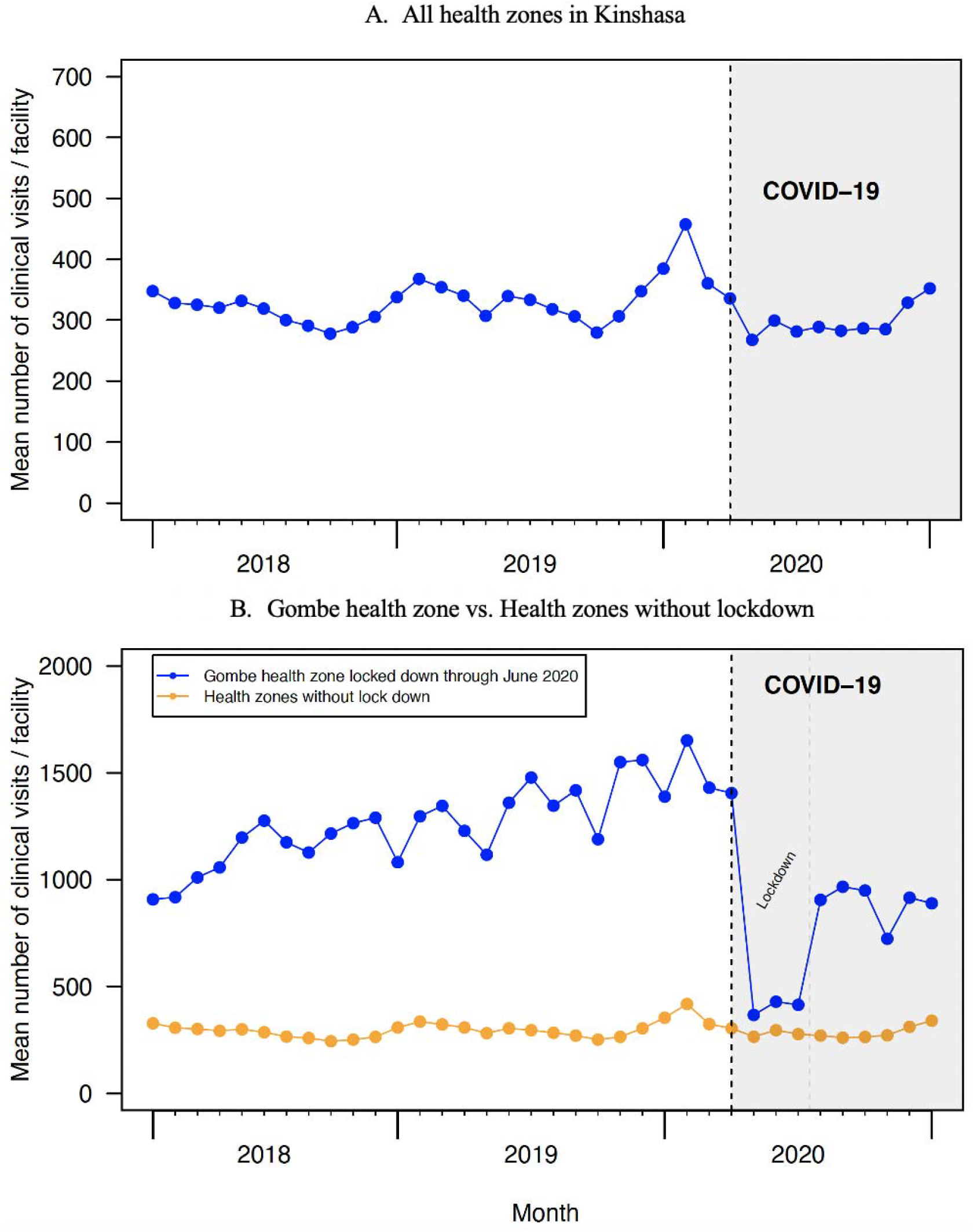
Time series of the mean number of total clinic visits

### Common infectious diseases

The start of the pandemic was associated with an immediate decrease in rates of visits for common infectious diseases, ranging from 24% reduction in visits for malaria (IRR: 0.76, 95% CI: 0.73–0.79) to 30% for pneumonia visits (IRR: 0.70, 95% CI: 0.65 –0.74) (Table 1). Important drops in RDTs were also observed. However, the trends of rates of visits for infectious diseases increased over time marginally, but significantly, following the start of the pandemic (Table 1). Our stratified analyses showed that, overall, decreases in rates of visits for malaria, diarrhea, and pneumonia were substantially larger in Gombe relative to the other health zones (Table 2, and Figure 4). In particular, in Gombe, clinic visits decreased immediately by 90% for malaria (IRR: 0.10, 95% CI: 0.05–0.20), 81% for diarrhea (IRR: 0.19, 95% CI: 0.08-0.43), and 75% for pneumonia (IRR: 0.25, 95% CI: 0.09–0.69). However, the use of health services also immediately increased following the end of the lockdown by ranging from approximately 60% for pneumonia to 75% for diarrhea and RDTs increased by more than two-fold during the same time period (Table 2 and Figure 4). It should be noted that these increases remained lower than the pre-pandemic rate (Appendix Table 6). In the health zones without a lockdown, visits for these infectious diseases declined by about only 20-30% immediately after onset of pandemic and stayed similar in the post-lockdown pandemic period (Table 2 and Figure 4).

**Figure 4.**
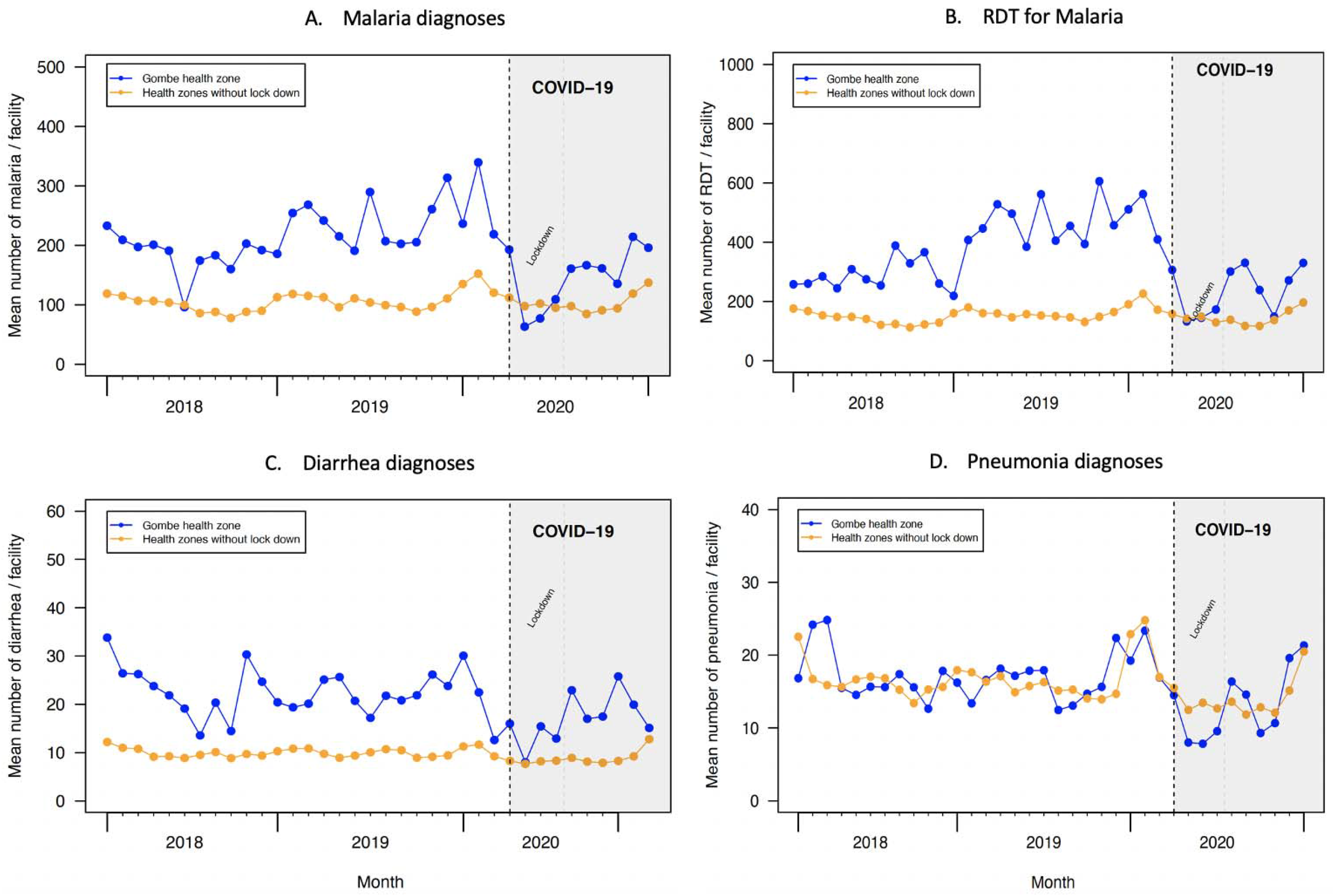
Time series of the mean number of (A) malaria diagnoses, (B) rapid diagnostic test for malaria, (C) diarrhea diagnoses and (D) pneumonia diagnoses, Gombe in comparison to other Health zones without a lockdown, Kinshasa, 2018-2020

### Non-communicable diseases

The pandemic was associated with an immediate decline in rates of visits for new diagnosis of NCDs: a 39% drop in for diabetes (IRR: 0.61, 95% CI: 0.55–0.66) and 16% drop for hypertension (IRR: 0.84, 95% CI: 0.79–0.89) were observed (Table 1). Stratified analysis showed that in Gombe, immediately following the COVID-19 and lockdown policy, visits decreased by 93% (IRR: 0.07, 95% CI: 0.03–0.19) for new diabetes patients and 77% for new hypertension patients (IRR: 0.23, 95% CI: 0.07–0.70); however, in the post-lockdown pandemic period, visits immediately increased by more than 50% for both diabetes and hypertension, but they still remained about 35% lower than in the pre-pandemic period (see Table 2, Figure 5, and Appendix Table 6). In the health zones without a lockdown, visits declined by only 10% for diabetes (IRR level change: 0.91, 95% CI: 0.82–0.99) and 15% for hypertension (IRR: 0.84, 95% CI: 0.79–0.89) (see Table 2 and Figure 5) and stayed similar in the post-lockdown pandemic period (Table 2 and Figure 5).

**Figure 5.**
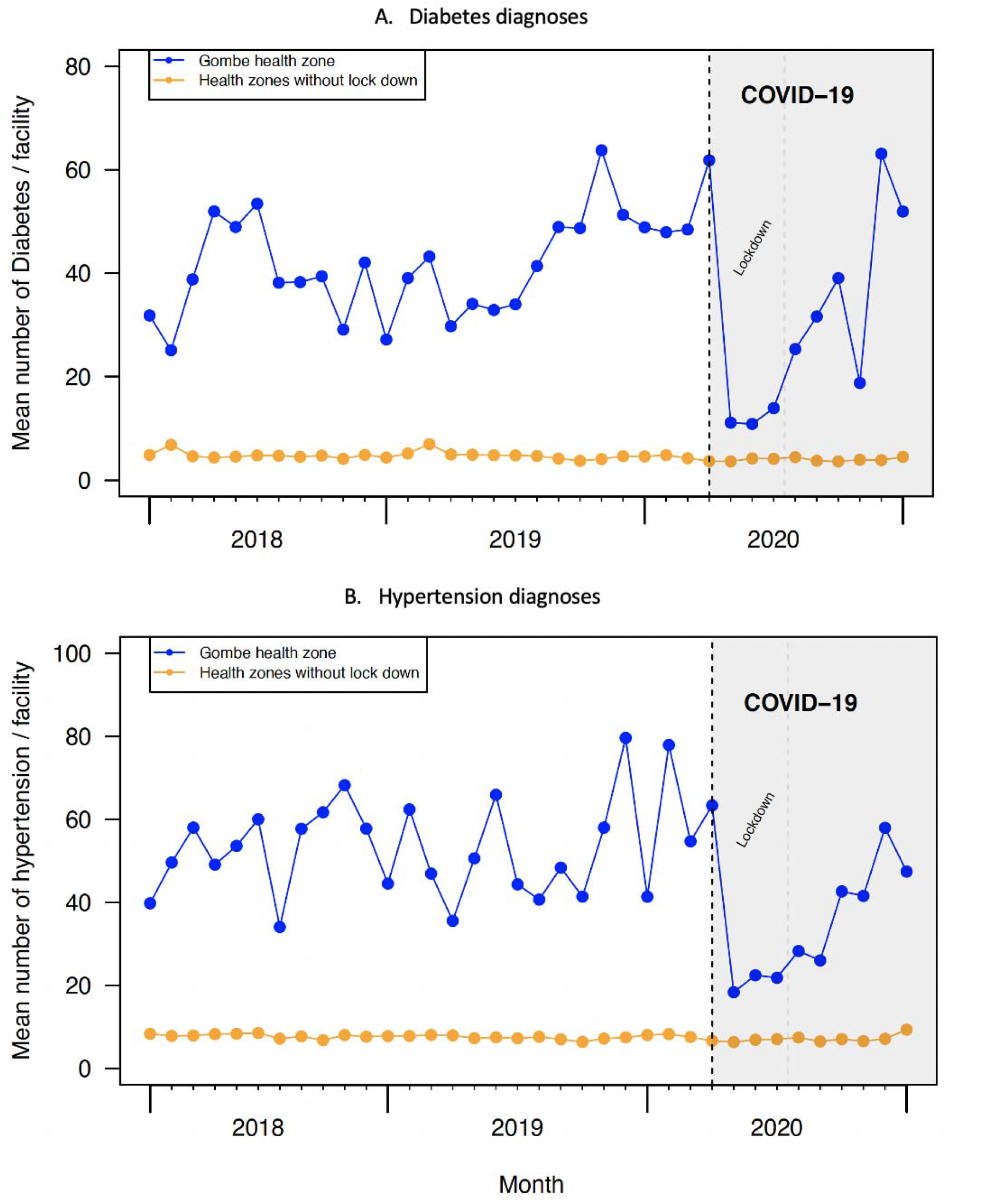
Time series of the mean number of new diagnoses (A) diabetes and (B) hypertension, Gombe in comparison to other health zones without a lockdown, Kinshasa, 2018-2020

### Maternal health

Unlike other health services, rates of first antenatal care visits, institutional deliveries, and second postnatal care visits increased modestly immediately following the start of pandemic (Table 1). However, the trend in rates of maternal health services did not increase significantly over time as compared with the trends that would have been expected without COVID-19 (Table 1). When examining changes in rates of the use of these maternal health services by lockdown policy, we found that overall rates of institutional deliveries and visits for second postnatal care were not substantially affected by the pandemic nor the lockdown policy (Table 2, Figure 6B and Appendix Figure 3). However, the rate of visits for antenatal care decreased by ∼45% in Gombe (IRR: 0.57, 95% CI: 0.35–0.91) immediately following the start of the pandemic and lockdown policy (Table 2 and Figure 6A).

**Figure 6.**
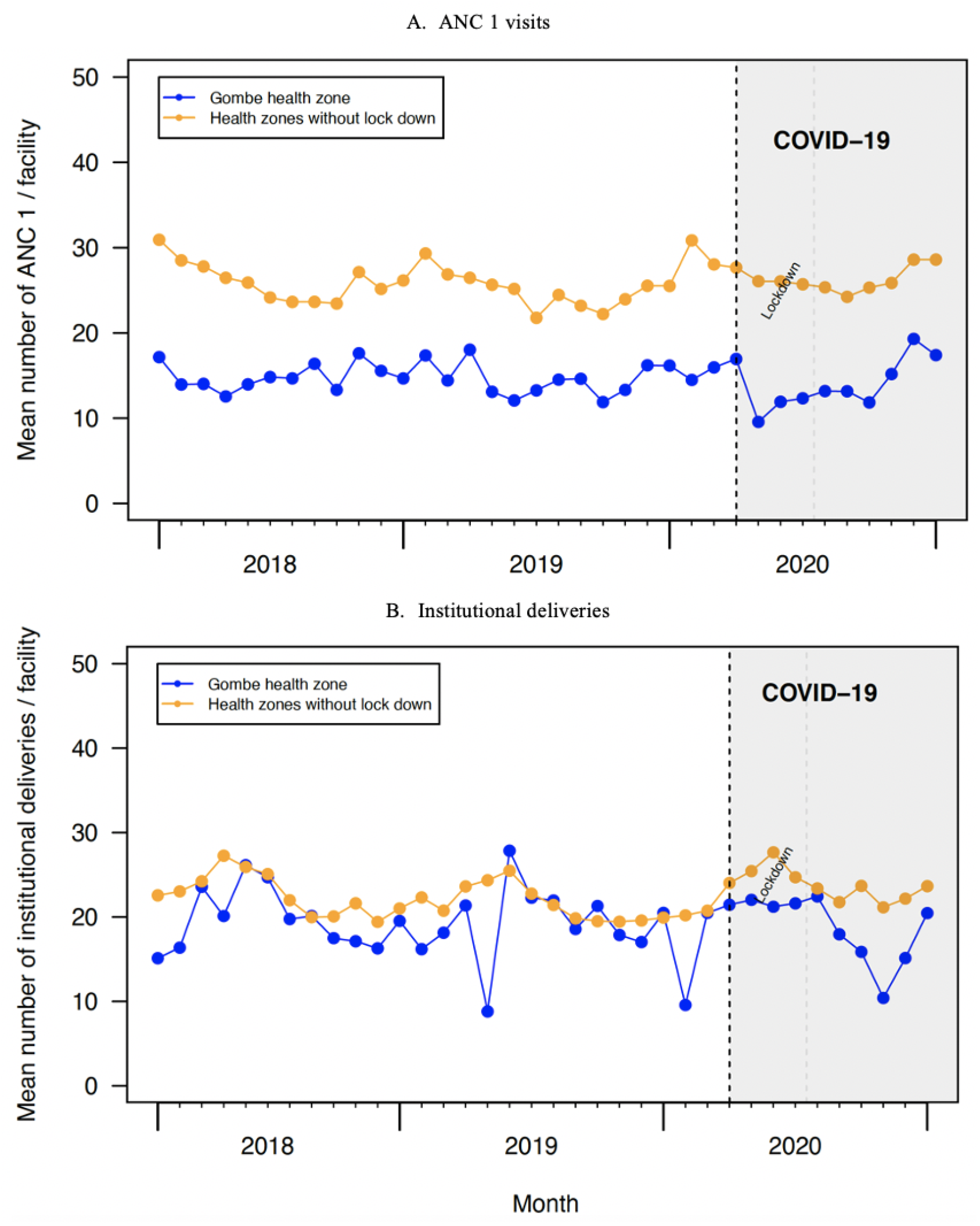
Time series of the mean number of (A) visits for first antenatal care and (B) institutional deliveries, Gombe in comparison to other health zones without a lockdown, Kinshasa, 2018-2020

### Vaccinations

Vaccinations were largely not affected by COVID-19 in Kinshasa (Table 1). Given few facilities (n=2) in the Gombe health zone reported vaccinations consistently during the study period, we were not able to perform additional subgroup analyses to understand the effect of the lockdown policy on vaccinations in the Gombe health zone.

## DISCUSSION

We found that the use of health services immediately decreased following the start of the COVID-19 pandemic in Kinshasa. Sizeable declines were observed for total clinic visits, as well as for visits for common infectious diseases (pneumonia, diarrhoea, malaria) and for new diagnoses of NCDs (diabetes and hypertension). While substantial reductions were recorded in Gombe (e.g., ∼83% for total visits), reductions were much less pronounced in other areas of Kinshasa (e.g., ∼16% for total visits). Following the full termination of the lockdown policy, total clinic visits, as well as visits for infectious diseases and NCDs, increased immediately by about two-fold; however, this increase was not sustained over time in most cases utilisation rates did not return to pre-pandemic levels. Hospitals were more affected than health centres and the use of maternal health services and vaccinations were considerably less affected.

Our study has several limitations. First, while we had a relatively long period of data before COVID-19, the periods during the lockdown included only three data points. While three data points are the minimum recommended for ITS studies, this rather short period may have affected the power of our study. Second, our analyses did not include all health facilities in Kinshasa. Specifically, we excluded health posts and other health facilities due to low reporting rates, as well as private health facilities. Similarly, several health centres and hospitals in Kinshasa were not included in our analyses because of inconsistent reporting of data. Finally, the Provincial Hospital of Kinshasa, the largest hospital in the city and that provides services to the whole city, is located Gombe. Hence, the lockdown of the Gombe area also affected the use of health services outside of Gombe, which we could not isolate from the effect of the lockdown. However, ITS does not require a comparison area thus the results are still consistent for each geographical region.

Although we found important reductions in the use of health services, we did not observe decreases as large as those predicted by modelling studies at the outset of the pandemic, with the exception of in Gombe. However, we also found that health service utilization did not return to pre-pandemic levels in the post-lockdown pandemic period, suggesting that short-run policies can also have longer-run effects.

While our study has shed light into changes in the use of health services in Kinshasa, we could not attribute these changes to specific mechanisms. From a policy perspective, it is important to determine if the observed changes were due mainly to change in the availability or accessibility of services as opposed to the demand for health services, including any impact factors such as fear or financial access.^14^ The fact that hospitals, where COVID-19 treatment centers had been located, saw bigger declines, might be evidence that fear of infection may have made people reluctant to visit such facilities but we did not observe a shift to health centers. This suggests that the population may have avoided visits to any health facility during the early pandemic period. In Kinshasa, COVID-19 was a stigmatizing disease and as fever and respiratory symptoms are common reasons for consultations, people may have preferred to stay at home and practice self-medication rather than risk getting diagnosed as a case. Whether there was an important shift to the private sector needs to be further assessed. However, there were no reports of overloaded private structures during this period. Most maternal and child health services tend to be delivered at health centres, which could also explain the smaller effects observed for these indicators.

Early in the pandemic, in Kinshasa there was widespread awareness that comorbidities with NCDs could exacerbate COVID-19 outcomes, which could partially explain the very large drops for these health services. The impact of other measures not directly affecting Kinshasa could have also played a role, for example, the decline in availability of RDTs for malaria due to international travel restrictions for several months. Complementary qualitative studies evaluating the perception of the population vis-à-vis health service use during an epidemic, but also evaluating the economic impact on household level of a pandemic with concurrent lockdown measures, should be investigated.

The COVID-19 pandemic has led to substantial health, economic and social effects globally. In this study, we document important declines in the use of health services during the first wave of the pandemic in Kinshasa, mostly notably in the parts of the city that were subject to lockdown. Given the relatively low numbers of cases and deaths recorded in Kinshasa during this time period, the effectiveness of such policies in containing COVID-19 needs to be weighed against its potential impact on population health. Integrating health information system data analysis with social sciences evidence can contribute to a comprehensive interpretation of the data.^36^

## Data Availability

The data are not publicly available.

## Acknowledgments

The findings of this study have been previously presented at the 2021 DHIS2 Annual Conference and the 2021 CUGH Annual Global Health Conference.

## SUPPLEMENTAL MATERIALS

### Statistical Appendix

We first attempted to fit segmented Poison mixed effect models, but we subsequently added a scale parameter (the Pearson X^2^ statistic divided by the residual degrees of freedom) to our regression models to adjust for overdispersion—i.e., the variance in the outcome measures exceeds the mean, violating the Poisson assumption. Our interrupted time series analysis model had the following basic general form for each outcome

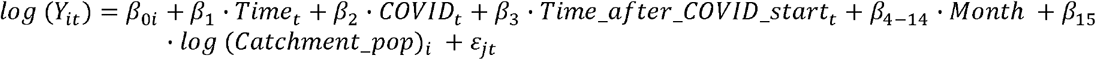

Where *Y*_*it*_ represents each of 14 indicators of health service use included in this study (e.g., total visits and ANC 1); subscripts *i* and *t* denote a health facility *i* at time *t. β* _0*i*_ represents the intercept of our model with both a fixed effect and facility-level random effects to account for heterogeneity in the volume of health service utilization. *Time* is the month at time *t* (running from January 2018 through December 2020, excluding March 2020). *COVID* indicates whether the COVID-19 pandemic (and response) had occurred at time *t*; and *time_after_COVID_start* represents the time *t* it was after the start of the pandemic (and response). *Month* denotes a dummy variable indexing month of the year using the month of January as the reference category to adjust for seasonality in services use. *Catchment_pop* is the estimated facility catchment population; we included the log of each facility catchment population as an offset in our model given the number of visits in a given facility depends on the size of the catchment population. In each model, the coefficients of interest are *ß*_*2*_, which indicates any immediate change in the rate of service use following the start of the pandemic (and response), and *ß*_*3*_, which indicates any change in the monthly trend in the rate of service use after the beginning of the pandemic (and response).

For the Gombe health zone, we run models that included segments (level and trend change) for lockdown period (April-June 2020) and for the period post lockdown (July-December 2020), allowing us to also estimate the impact of stopping the lockdown policy. These models had the following basic general form for each outcome:

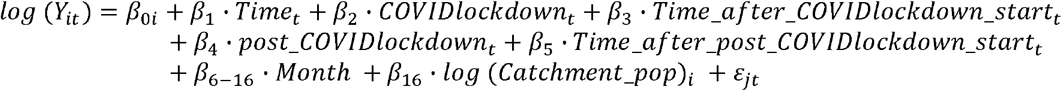

Where *Y*_*it*_ represents each of 14 indicators of health service use included in this study, subscripts *i* and *t* denote a health facility *i* at time *t. β* _0*i*_ represents the intercept of our model with both a fixed effect and facility-level random effects to account for heterogeneity in the volume of health service utilization. *Time* is the month at time *t* (running from January 2018 through December 2020, excluding March 2020). *COVIDlockdown* indicates whether the COVID-19 pandemic and lockdown had occurred at time *t*; and *time_after_COVIDlockdown_start* represents the time *t* it was after the start of the pandemic and lockdown. *Post_COVIDlockdown* indicates whether the lockdown had been lifted at time *t*; and *time_after_post_COVIDlockdown_start* represents the time *t* it was after the lockdown was lifted. *Month* denotes a dummy variable indexing month of the year using the month of January as the reference category to adjust for seasonality in services use. *Catchment_pop* is the estimated facility catchment population; we included the log of each facility catchment population as an offset in our model given the number of visits in a given facility depends on the size of the catchment population. In each model run for the Gombe health zone, the coefficients of interest are *ß*_*2*_, which indicates any immediate change in the rate of service use following the start of the pandemic and lockdown; *ß*_*3*_, which indicates any change in the monthly trend in the rate of service use after the beginning of the pandemic and lockdown; *ß*_*4*_, which indicates any immediate change in the rate of service use following the stop of start of the lockdown; and *ß*_*5*_, which indicates any change in the monthly trend in the rate of service use after the lockdown was lifted.

Additionally, to minimize potential overestimation of the impact of COVID-19 and related response measures on services use, especially pneumonia, models for total clinic visits and visits for common infectious diseases included a dummy variable to adjust for a pneumonia outbreak that took place in Kinshasa from December 2019 to February 2020. Lastly, to assess whether the post-lockdown rates in service use returned to pre-pandemic rates, we run segmented Poison mixed effects models with two segments: pre-pandemic (January 2018-February 2020) and post-lockdown (July-December 2020).

**Appendix Table 1.** Overall reporting rate for indicators included in this study.

| Indicators | Mean reporting rate, % | Median reporting rate, % | Minimum reporting rate, % | Maximum reporting rate, % |
| --- | --- | --- | --- | --- |
| Clinic visits | 87.96 | 100 | 3.84 | 100 |
| Uncomplicated malaria case diagnosis | 83.64 | 92.30 | 3.84 | 100 |
| First antenatal care | **81.88** | **92.30** | 3.84 | 100 |
| RDT for malaria done | **81.11** | **90.00** | **3. 3.84** | 100 |
| Institutional deliveries | 79.00 | 90.00 | 3.84 | 100 |
| Second postnatal care | 70.99 | 80.76 | 3.84 | 100 |
| OPV 1 | 65.53 | 76.92 | 3.84 | 100 |
| DTC-HepB Hib1 | 65.42 | 80.00 | 3.84 | 100 |
| PCV-13 (1) | 64.81 | 76.92 | 3.84 | 100 |
| BCG | 62.28 | 73.07 | 3.84 | 100 |
| Uncomplicated diarrhea case diagnosis | 61.04 | 69.23 | 3.84 | 100 |
| Hypertension diagnosis | 56.62 | 61.53 | 3.84 | 100 |
| Uncomplicated pneumonia case diagnosis | 49.10 | 48.07 | 3.84 | 100 |
| Diabetes diagnosis | 43.13 | 38.46 | 3.84 | 100 |

These are reporting rates before exclusion of facilities that did not meet in our inclusion criteria discussed earlier in the Methods Section.

**Appendix Table 2.** Parameter estimates of the overall effect of COVID-19 outbreak and related response measures on rates of total clinic visits and visits for common infectious diseases when the pneumonia outbreak was not adjusted for.

| Indicators | Level change |  | Trend change |  |
| --- | --- | --- | --- | --- |
|  | IRR (95% CI) | p-value | IRR (95% CI) | p-value |
| **Overall visits** |  |  |  |  |
| Total clinic visits | 0.74 (0.72, 0.76) | <0.001 | 1.008 (1.004, 1.013) | <0.001 |
| **Common IDs** |  |  |  |  |
| Malaria | 0.76 (0.73, 0.79) | <0.001 | 1.01 (1.00, 1.02) | <0.001 |
| RDT | 0.71 (0.68, 0.74) | <0.001 | 1.01 (1.00, 1.02) | <0.001 |
| Diarrhea | 0.76 (0.72, 0.80) | <0.001 | 1.01 (1.01, 1.03) | <0.001 |
| Pneumonia | 0.66 (0.62, 0.71) | <0.001 | 1.02 (1.01, 1.03) | <0.001 |

IRR: incidence rate ratio; CI: confidence interval; IDs, infectious diseases; RDT, rapid diagnostic test for malaria

**Appendix Table 3.** Facilities reported through DHIS2 in Kinshasa for each indicator and number of health facilities included in the study main analytic sample.

|  | Health Centre | Hospital |
| --- | --- | --- |
| All Facilities reported, N | 854 | 121 |
| Facilities included for each indicator, n (%) |  |  |
| Total clinic visits | 747 (87.47) | 106 (87.60) |
| Malaria | 720 (84.30) | 102 (84.29) |
| RDT | 704 (82.43) | 99 (81.81) |
| Diarrhea | 608 (71.19) | 89 (73.55) |
| Pneumonia | 374 (43.79) | 78 (64.46) |
| ANC 1 | 677 (79.27) | 92 (76.03) |
| Delivery | 643 (75.29) | 94 (77.68) |
| PNC 2 | 578 (67.68) | 84 (69.42) |
| DTP1 | 446 (52.22) | 3 (2.47) |
| BCG | 439 (51.40) | 2 (1.65) |
| OPV | 436 (51.03) | 2 (1.65) |
| PVC 13 | 442 (51.75) | 3 (2.47) |
| Diabetes | 388 (45.43) | 87 (71.90) |
| Hypertension | 654 (76.58) | 101 (83.47) |

RDT, rapid diagnostic test for malaria; ANC, antenatal care; PNC, postnatal care; DTP: diphtheria, pertussis, and tetanus vaccine; BCG, Bacillus Calmette–Guérin vaccine; OPV, Oral poliovirus vaccine; PCV-13, Pneumococcal Conjugate Vaccine 13.

**Appendix Table 4.** Volume of services included in our analysis.

| Indicators | Volume of services over the study period | Average per facility per month | Standard deviation | Median | Minimum | Maximum |
| --- | --- | --- | --- | --- | --- | --- |
| Overall clinic visits | 9158657 | 321.89 | 37.19 | 319.75 | 267.74 | 457.42 |
| Infectious diseases |  |  |  |  |  |  |
| Malaria | 2879344 | 106.76 | 15.97 | 102.36 | 80.74 | 159.27 |
| RDT | 3938939 | 156.20 | 24.58 | 156.50 | 120.79 | 238.59 |
| Diarrhea | 200433 | 9.85 | 1.20 | 9.64 | 7.68 | 12.77 |
| Pneumonia | 203113 | 15.79 | 2.89 | 15.45 | 11.90 | 24.54 |
| NCDs |  |  |  |  |  |  |
| Diabetes | 69761 | 6.27 | 1.10 | 6.50 | 3.78 | 8.52 |
| Hypertension | 186644 | 8.74 | 1.02 | 8.88 | 6.48 | 10.55 |
| Maternal health services |  |  |  |  |  |  |
| ANC 1 | 637321 | 25.54 | 2.09 | 25.37 | 21.52 | 30.49 |
| Facility deliveries | 522932 | 22.36 | 2.35 | 21.99 | 19.25 | 27.52 |
| PNC 2 | 320670 | 16.45 | 1.78 | 16.32 | 13.93 | 19.74 |
| Vaccinations |  |  |  |  |  |  |
| DTP 1 | 477888 | 36.87 | 1.61 | 36.46 | 34.48 | 40.85 |
| BCG | 540590 | 42.80 | 3.07 | 42.18 | 37.08 | 53.46 |
| OPV | 493559 | 40.35 | 2.44 | 40.13 | 35.63 | 45.61 |
| PVC 13 | 474753 | 37.24 | 1.52 | 37.24 | 34.54 | 41.13 |

RDT, rapid diagnostic test for malaria; ANC, antenatal care; PNC, postnatal care; DTP: diphtheria, pertussis, and tetanus vaccine; BCG, Bacillus Calmette–Guérin vaccine; OPV, Oral poliovirus vaccine; PCV-13, Pneumococcal Conjugate Vaccine 13; NCDs, non-communicable diseases.

**Appendix Table 5.**
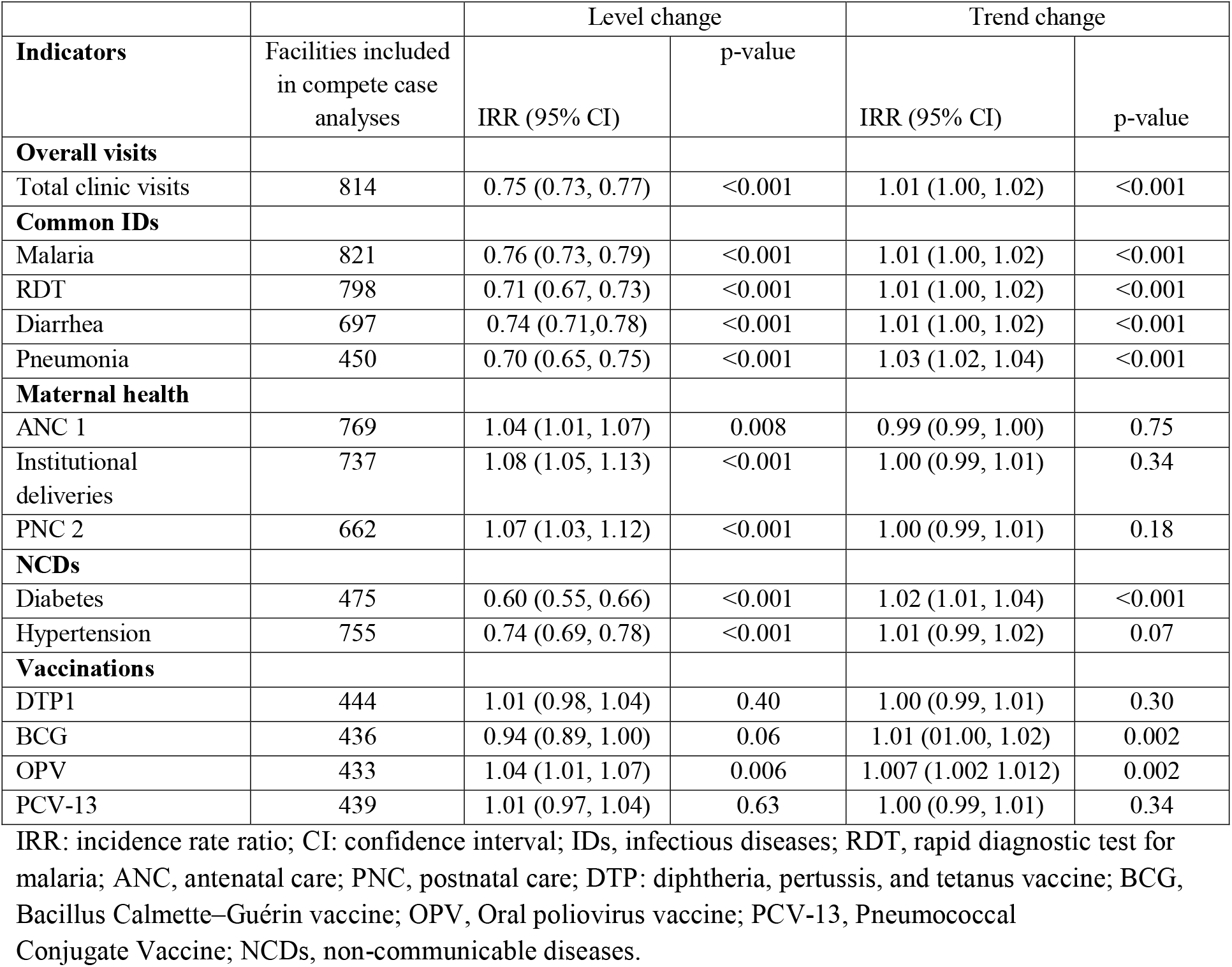
Complete case analyses. Parameter estimates of the overall effect of COVID-19 outbreak and related response measures on rates of total clinic visits, visits for common infectious diseases and non-communicable diseases, institutional deliveries, and vaccinations in Kinshasa, DRC.

| Indicators | Facilities included in complete case analyses | Level change |  | Trend change |  |
| --- | --- | --- | --- | --- | --- |
|  |  | IRR (95% CI) | p-value | IRR (95% CI) | p-value |
| **Overall visits** |  |  |  |  |  |
| Total clinic visits | 814 | 0.75 (0.73, 0.77) | <0.001 | 1.01 (1.00, 1.02) | <0.001 |
| **Common IDs** |  |  |  |  |  |
| Malaria | 821 | 0.76 (0.73, 0.79) | <0.001 | 1.01 (1.00, 1.02) | <0.001 |
| RDT | 798 | 0.71 (0.67, 0.73) | <0.001 | 1.01 (1.00, 1.02) | <0.001 |
| Diarrhea | 697 | 0.74 (0.71, 0.78) | <0.001 | 1.01 (1.00, 1.02) | <0.001 |
| Pneumonia | 450 | 0.70 (0.65, 0.75) | <0.001 | 1.03 (1.02, 1.04) | <0.001 |
| **Maternal health** |  |  |  |  |  |
| ANC 1 | 769 | 1.04 (1.01, 1.07) | 0.008 | 0.99 (0.99, 1.00) | 0.75 |
| Institutional deliveries | 737 | 1.08 (1.05, 1.13) | <0.001 | 1.00 (0.99, 1.01) | 0.34 |
| PNC 2 | 662 | 1.07 (1.03, 1.12) | <0.001 | 1.00 (0.99, 1.01) | 0.18 |
| **NCDs** |  |  |  |  |  |
| Diabetes | 475 | 0.60 (0.55, 0.66) | <0.001 | 1.02 (1.01, 1.04) | <0.001 |
| Hypertension | 755 | 0.74 (0.69, 0.78) | <0.001 | 1.01 (0.99, 1.02) | 0.07 |
| **Vaccinations** |  |  |  |  |  |
| DTP1 | 444 | 1.01 (0.98, 1.04) | 0.40 | 1.00 (0.99, 1.01) | 0.30 |
| BCG | 436 | 0.94 (0.89, 1.00) | 0.06 | 1.01 (1.00, 1.02) | 0.002 |
| OPV | 433 | 1.04 (1.01, 1.07) | 0.006 | 1.007 (1.002, 1.012) | 0.002 |
| PCV-13 | 439 | 1.01 (0.97, 1.04) | 0.63 | 1.00 (0.99, 1.01) | 0.34 |

IRR: incidence rate ratio; CI: confidence interval; IDs, infectious diseases; RDT, rapid diagnostic test for malaria; ANC, antenatal care; PNC, postnatal care; DTP: diphtheria, pertussis, and tetanus vaccine; BCG, Bacillus Calmette–Guérin vaccine; OPV, Oral poliovirus vaccine; PCV-13, Pneumococcal Conjugate Vaccine; NCDs, non-communicable diseases.

**Appendix Table 6.**
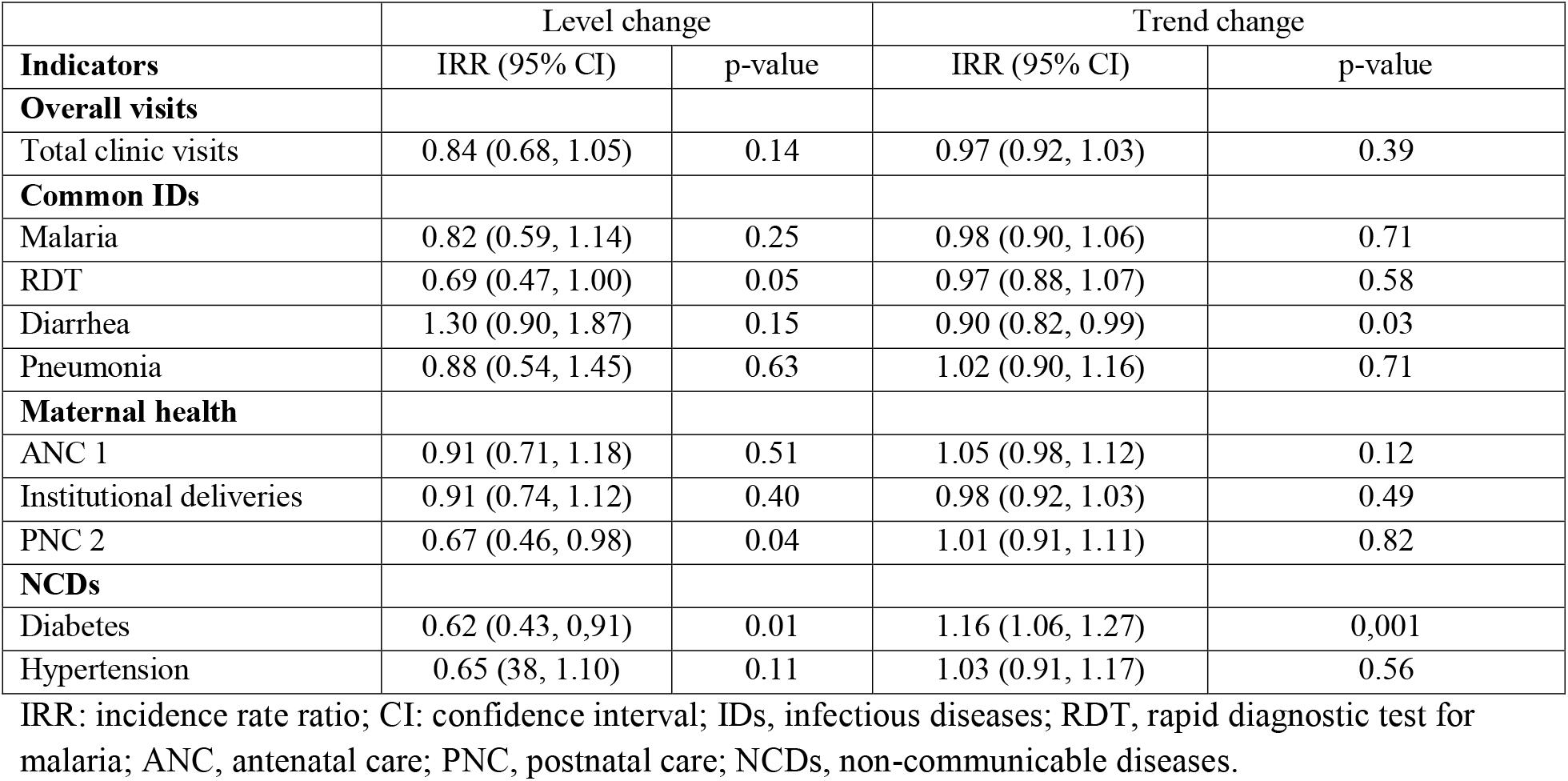
Parameters estimates comparing pre-pandemic and post lockdown periods in the Gombe health zone, DRC

| **Indicators** | **Level change** |  | **Trend change** |  |
| --- | --- | --- | --- | --- |
|  | **IRR (95% CI)** | **p-value** | **IRR (95% CI)** | **p-value** |
| **Overall visits** |  |  |  |  |
| Total clinic visits | 0.84 (0.68, 1.05) | 0.14 | 0.97 (0.92, 1.03) | 0.39 |
| **Common IDs** |  |  |  |  |
| Malaria | 0.82 (0.59, 1.14) | 0.25 | 0.98 (0.90, 1.06) | 0.71 |
| RDT | 0.69 (0.47, 1.00) | 0.05 | 0.97 (0.88, 1.07) | 0.58 |
| Diarrhea | 1.30 (0.90, 1.87) | 0.15 | 0.90 (0.82, 0.99) | 0.03 |
| Pneumonia | 0.88 (0.54, 1.45) | 0.63 | 1.02 (0.90, 1.16) | 0.71 |
| **Maternal health** |  |  |  |  |
| ANC 1 | 0.91 (0.71, 1.18) | 0.51 | 1.05 (0.98, 1.12) | 0.12 |
| Institutional deliveries | 0.91 (0.74, 1.12) | 0.40 | 0.98 (0.92, 1.03) | 0.49 |
| PNC 2 | 0.67 (0.46, 0.98) | 0.04 | 1.01 (0.91, 1.11) | 0.82 |
| **NCDs** |  |  |  |  |
| Diabetes | 0.62 (0.43, 0.91) | 0.01 | 1.16 (1.06, 1.27) | 0.001 |
| Hypertension | 0.65 (0.38, 1.10) | 0.11 | 1.03 (0.91, 1.17) | 0.56 |

IRR: incidence rate ratio; CI: confidence interval; IDs, infectious diseases; RDT, rapid diagnostic test for malaria; ANC, antenatal care; PNC, postnatal care; NCDs, non-communicable diseases.

**Appendix Figure 1.**
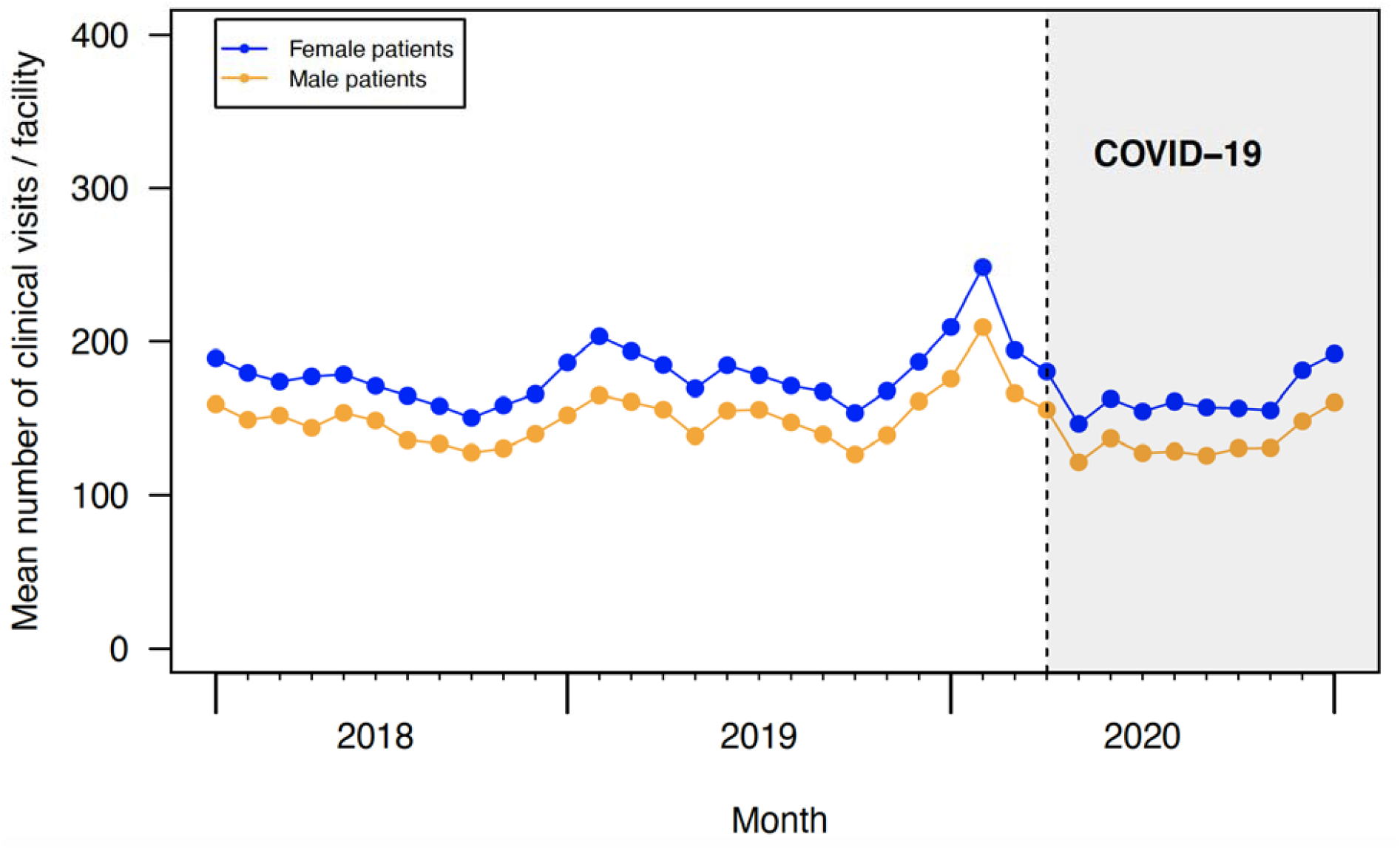
Time series of the mean number of clinic visits stratified by female and male patients across Kinshasa.

**Appendix Figure 2.**
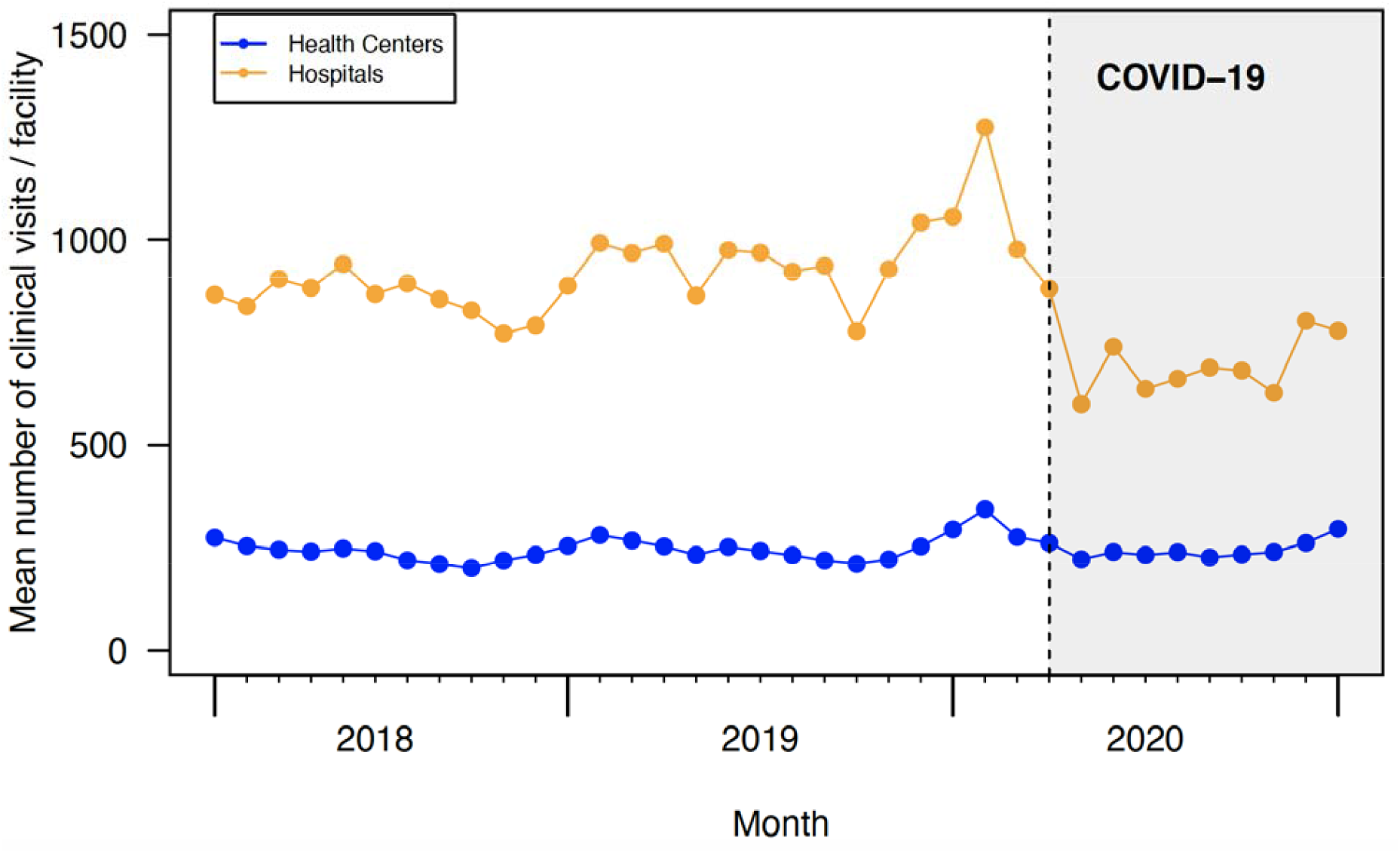
Time series of the mean number of clinic visits by health centers and hospitals.

**Appendix Figure 3.**
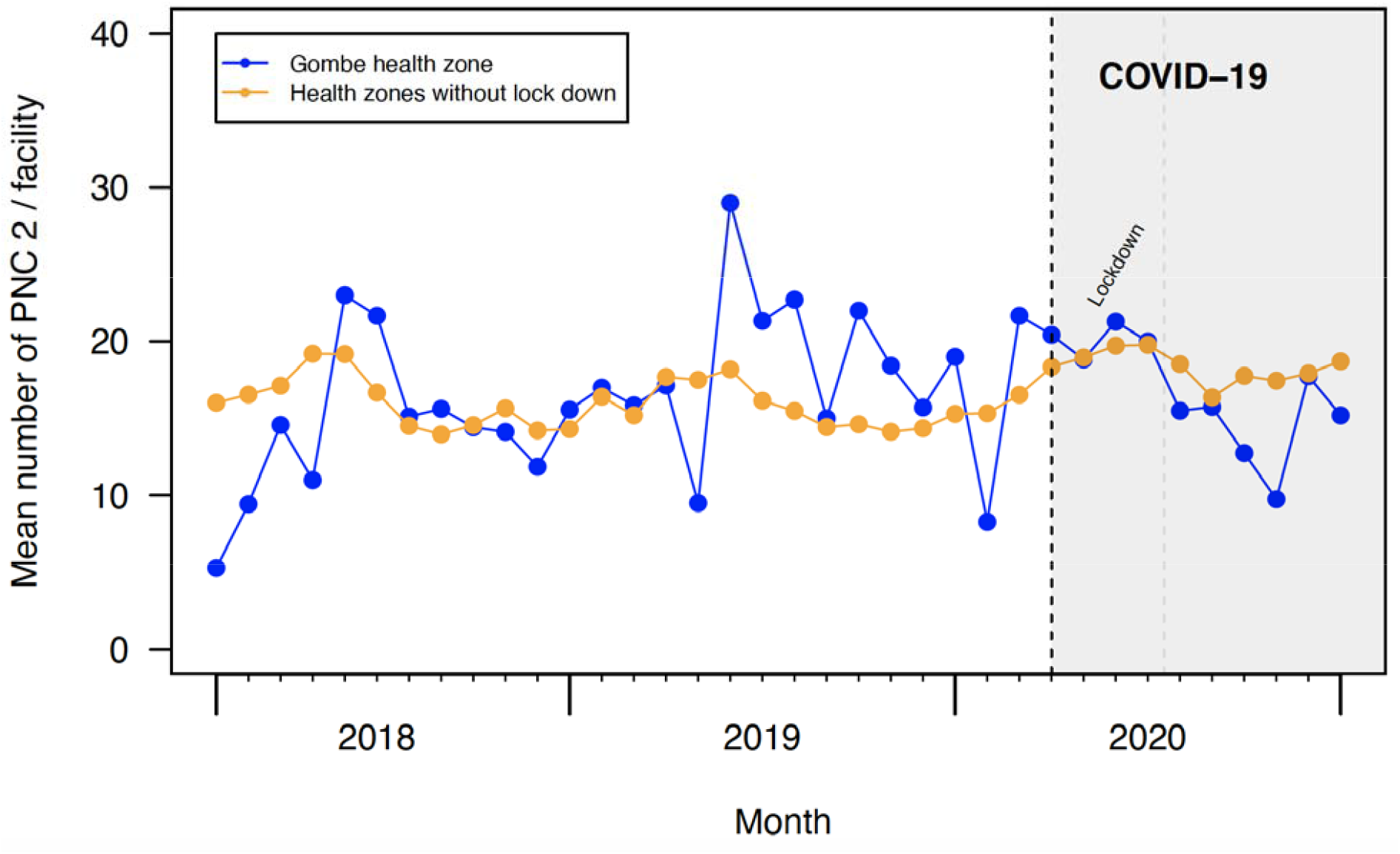
Time series of the mean number of visits for second postnatal care

## REFERENCES

1 Cabore JW, Karamagi HC, Kipruto H, et al. The potential effects of widespread community transmission of SARS-CoV-2 infection in the World Health Organization African Region: a predictive model. Bmj Global Heal 2020; 5: e002647.

2 WHO. WHO Coronavirus Disease (COVID-19) Dashboard. undefined. https://covid19.who.int/ (accessed Feb 11, 2021).

3 Wilhelm JA, Helleringer S. Utilization of non-Ebola health care services during Ebola outbreaks: a systematic review and meta-analysis. J Glob Health 2019; 9: 010406.

4 Elston JWT, Cartwright C, Ndumbi P, Wright J. The health impact of the 2014–15 Ebola outbreak. Public Health 2017; 143: 60–70.

5 Helleringer S, Noymer A. Magnitude of Ebola relative to other causes of death in Liberia, Sierra Leone, and Guinea. Lancet Global Heal 2015; 3: e255.6.

6 Walker PGT, White MT, Griffin JT, Reynolds A, Ferguson NM, Ghani AC. Malaria morbidity and mortality in Ebola-affected countries caused by decreased health-care capacity, and the potential effect of mitigation strategies: a modelling analysis. Lancet Infect Dis 2015; 15: 825–32.

7 Sochas L, Channon AA, Nam S. Counting indirect crisis-related deaths in the context of a low-resilience health system: the case of maternal and neonatal health during the Ebola epidemic in Sierra Leone. Health Policy Plann 2017; 32: iii32–9.

8 Takahashi S, Metcalf CJE, Ferrari MJ, et al. Reduced vaccination and the risk of measles and other childhood infections post-Ebola. Science 2015; 347: 1240–2.

9 Plucinski MM, Guilavogui T, Sidikiba S, et al. Effect of the Ebola-virus-disease epidemic on malaria case management in Guinea, 2014: a cross-sectional survey of health facilities. Lancet Infect Dis 2015; 15: 1017 1023.

10 Tattevin P, Baysah MK, Raguin G, et al. Retention in care for HIV-infected patients in the eye of the Ebola storm: lessons from Monrovia, Liberia. Aids 2015; 29: N1 2.

11 Leuenberger D, Hebelamou J, Strahm S, et al. Impact of the Ebola epidemic on general and HIV care in Macenta, Forest Guinea, 2014. Aids Lond Engl 2015; 29: 1883–7.

12 Evans DK, Goldstein M, Popova A. Health-care worker mortality and the legacy of the Ebola epidemic. Lancet Global Heal 2015; 3: e439–40.

13 McLean KE, Abramowitz SA, Ball JD, et al. Community-based reports of morbidity, mortality, and health-seeking behaviours in four Monrovia communities during the West African Ebola epidemic. Glob Public Health 2016; 13: 1–17.

14 Morse B, Grépin KA, Blair RA, Tsai L. Patterns of demand for non-Ebola health services during and after the Ebola outbreak: panel survey evidence from Monrovia, Liberia. Bmj Global Heal 2016; 1: e000007.

15 Delamou A, Ayadi AME, Sidibe S, et al. Effect of Ebola virus disease on maternal and child health services in Guinea: a retrospective observational cohort study. Lancet Global Heal 2017; 5. DOI:10.1016/s2214-109x(17)30078-5.

16 Camara BS, Delamou A, Diro E, et al. Effect of the 2014/2015 Ebola outbreak on reproductive health services in a rural district of Guinea: an ecological study. T Roy Soc Trop Med H 2017; 111: 1–8.

17 Wagenaar BH, Augusto O, Beste J, et al. The 2014–2015 Ebola virus disease outbreak and primary healthcare delivery in Liberia: Time-series analyses for 2010–2016. Plos Med 2018; 15: e1002508.

18 Roberton T, Carter ED, Chou VB, et al. Early estimates of the indirect effects of the COVID-19 pandemic on maternal and child mortality in low-income and middle-income countries: a modelling study. Lancet Global Heal 2020; 8: e901–8.

19 Hogan AB, Jewell BL, Sherrard-Smith E, et al. Potential impact of the COVID-19 pandemic on HIV, tuberculosis, and malaria in low-income and middle-income countries: a modelling study. Lancet Global Heal 2020. DOI:10.1016/s2214-109x(20)30288-6.

20 WHO. The potential impact of health service disruptions on the burden of malaria. undefined 2020; published online April 23. https://www.who.int/publications/i/item/9789240004641.

21 Sherrard-Smith E, Hogan AB, Hamlet A, et al. The potential public health consequences of COVID-19 on malaria in Africa. Nat Med 2020; 26: 1411–6.

22 Press WH, Levin RC. Modeling, post COVID-19. Science 2020; 370: 1015–1015.

23 Siedner MJ, Kraemer JD, Meyer MJ, et al. Access to primary healthcare during lockdown measures for COVID-19 in rural South Africa: an interrupted time series analysis. Bmj Open 2020; 10: e043763.

24 Chandir S, Siddiqi DA, Setayesh H, Khan AJ. Impact of COVID-19 lockdown on routine immunisation in Karachi, Pakistan. Lancet Global Heal 2020. DOI:10.1016/s2214-109x(20)30290-4.

25 Kc A, Gurung R, Kinney MV, et al. Effect of the COVID-19 pandemic response on intrapartum care, stillbirth, and neonatal mortality outcomes in Nepal: a prospective observational study. Lancet Global Heal 2020. DOI:10.1016/s2214-109x(20)30345-4.

26 Nachega JB, Mbala-Kingebeni P, Otshudiema J, Zumla A, Tam-Fum J-JM. The colliding epidemics of COVID-19, Ebola, and measles in the Democratic Republic of the Congo. Lancet Global Heal 2020; 8: e991–2.

27 Lal A, Ashworth HC, Dada S, Hoemeke L, Tambo E. Optimizing Pandemic Preparedness and Response Through Health Information Systems: Lessons Learned From Ebola to COVID-19. Disaster Med Public 2020; : 1–8.

28 Hung YW, Hoxha K, Irwin BR, Law MR, Grépin KA. Using Routine Health Information Data for Research in Low- and Middle-Income Countries: A Systematic Review. undefined 2020. DOI:10.21203/rs.3.rs-26377/v1.

29 Hung YW, Law MR, Cheng L, et al. Impact of a Free Health Care Policy in the Democratic Republic of the Congo During an Ebola Outbreak: An Interrupted Time-Series Analysis. Ssrn Electron J 2019. DOI:10.2139/ssrn.3420410.

30 Bruner B, Combet V, Callahan S, et al. The Role of the Private Sector in Improving the Performance of the Health System in the Democratic Republic of Congo. Bethesda, MD, 2018.

31 AFRO W. First case of COVID-19 confirmed in the Democratic Republic of the Congo. undefined. 2020; published online March 10. https://www.afro.who.int/news/first-case-covid-19-confirmed-democratic-republic-congo (accessed Feb 11, 2021).

32 Evaluation M. Mapping the Stages of MEASURE Evaluation’s Data Use Continuum to DHIS 2: An Example from the Democratic Republic of the Congo. Chapel Hill, NC, 2019.

33 Hung YW, Law MR, Cheng L, et al. Impact of a free care policy on the utilisation of health services during an Ebola outbreak in the Democratic Republic of Congo: an interrupted time-series analysis. Bmj Global Heal 2020; 5: e002119.

34 Wagner AK, Soumerai SB, Zhang F, Ross-Degnan D. Segmented regression analysis of interrupted time series studies in medication use research. J Clin Pharm Ther 2002; 27: 299 309.

35 Hategeka C, Ruton H, Karamouzian M, Lynd LD, Law MR. Use of interrupted time series methods in the evaluation of health system quality improvement interventions: a methodological systematic review. Bmj Global Heal 2020; 5: e003567.

36 Carter SE, Gobat N, Zambruni JP, et al. What questions we should be asking about COVID-19 in humanitarian settings: perspectives from the Social Sciences Analysis Cell in the Democratic Republic of the Congo. Bmj Global Heal 2020; 5: e003607.

## References for the Appendix

1. SECRETARIAT TECHNIQUE COVID-19. Le plan de préparation et de riposte contre l’épidémie au covid-19 en République Démocratique Du Congo. 2020.

2. Ministry of Health DRC. Directives prises par le Gouvernement, https://www.stopcoronavirusrdc.info/directives-prises-par-le-gouvernement (2021).

